# Growing up in poverty, growing old in frailty: The life course shaping of health in America, Britain, and Europe – a prospective and retrospective study

**DOI:** 10.1101/2024.03.07.24303906

**Authors:** Gindo Tampubolon

## Abstract

**Background:** Childhood poverty is directly associated with many health outcomes in late life irrespective of youth health and of variation in health systems. The childhood poor in America, Britain and Europe have reported worse cognitive, muscle and mental functions in their fifties to nineties. But it is not known whether they have higher probabilities of experiencing frailty as their childhood recollections are likely to be erroneous.

**Materials and methods:** Some 79428 adults aged 50 and older retrospectively recalled their childhood conditions at ten and underwent prospective examinations to construct their Fried’s frailty phenotype. Childhood conditions in ELSA and SHARE include number of books, number of rooms, number of people, presence of running hot or cold water, fixed bath, indoor lavatory and central heating. Across in America, these are mostly replaced with financial hardship indicators including having to move because of family debt. Childhood poverty is a latent construct of error-laced recollection and its distal fully adjusted association with frailty phenotype is estimated with fixed effects probit model.

**Results:** Childhood poverty associates with higher probabilities of being frail (0.1097 ± 0.0169, p < 0.001) in 29 countries of America, Britain and Europe. Furthermore, women have higher probabilities of being frail (0.3051 ± 0.0152, p < 0.001). Age, education, wealth, marital status and youth illness exert influences on the probabilities of being frail. Sensitivity analyses were conducted using random effects model and by stratifying on sex.

**Discussion:** Evidence is mounting that childhood can last a life time, affecting cognitive and muscle function, mental health and now frailty. This evidence calls for urgent actions to eliminate child poverty on account of its lifelong rewards. (271 + 4476 words)

## Introduction

Childhood poverty is a risk factor for old age disability, dysfunction and disease in America, Britain, China and Europe.(1–8) It raises the question whether childhood poverty also marks older adults’ frailty, a syndrome of systemic decline across multiple organ functions which often entails adverse clinical outcomes, high costs of care and mortality.(9,10) This question extends the term ‘long arm of childhood conditions,’ coined by Hayward and Gorman who studied the influence of childhood poverty on mortality in American males in their forties and fifties.(11) Large and ongoing ageing surveys have provided evidence to support the links across the life course. Twenty eight trans-Atlantic countries from America through Britain to Israel report life course links that persist, marking muscle function, depression status and cognitive function of older adults who grew up in poverty.(3) With such cross-country comparative design, any new evidence on life course shaping of frailty can be used to advance efforts during the UN Decade of Healthy Ageing.(12) Such efforts are urgent because frailty –typically though not invariably measured as frailty phenotype or index– reduces older adults’ wellbeing while imposing high costs on society.(9,13,14)

This work aims to test the hypothesis of life course shaping of frailty in America, Britain and Europe which posits that childhood poverty continues to mark old age frailty even after youth or adult conditions are considered – the lifelong association between poverty and frailty remains. To achieve this three sister studies (Health & Retirement Study – HRS, English Longitudinal Study of Ageing – ELSA, Survey of Health Ageing & Retirement in Europe – SHARE)(15–17) covering 29 countries are used in a cross-country fixed effect design following earlier studies.(3,4) To guide further steps the lens of social determinants of health as links across the life course is adopted, defined as “conditions in the environments in which people are born and *raised*, live, learn, work, play, worship, and *age* that affect a wide range of health, functioning, and quality-of-life outcomes”.(18,19) The investigation which posited the life course association studied midlife American males; it was silent on the experience of large sections of the population i.e. females or much older adults. Thus cross-country studies of both sexes constitute a specific test of the life course shaping of frailty, helping to fathom its reach and limits. In sum, how one was raised materially in countries of America, Britain and Europe can have lasting effects on the chance of experiencing frailty in old age.

When designing the test one encounters different concepts and indicators for childhood conditions in the empirical literature. Childhood poverty, childhood socioeconomic position and adverse childhood experience were some concepts used. These were captured as a scale or latent constructs with various indicators. These in turn vary including parental social class, education and employment self-report of family financial hardship or bankruptcy; being raised in a foster family; material facilities of childhood home such as number of rooms and people (indicating overcrowding), availability of books, central heating or indoor lavatory. This study does not use the adverse childhood experience scale for two reasons. First, the scale is complementary to the other concepts. Second, the scale has more psychological or subjective elements which suggests more susceptibility to bias. I am not aware of any test of the magnitude of the bias, for instance by comparing an independent report during childhood and during old age, unlike childhood poverty which has undergone such test.(1,2)

This work examines the life course shaping hypothesis, which if supported in rich ageing nations, means efforts to secure healthy ageing around the world can rightly consider the risk factor of childhood poverty, including in the [UN Decade of Healthy Ageing](https://www.who.int/initiatives/decade-of-healthy-ageing).(12)

## Methods and materials

### Outcome: frailty phenotype in HRS, ELSA and SHARE

The analytic sample was constructed from a family of ageing studies: HRS, ELSA and SHARE.(15–17) Already in 2015 Theo and colleagues found that there were 226 constructs of frailty phenotype.(20) I avoided devising another construct because replicability is a critical practice for cross-country study, choosing to follow closely studies of Fried, Bandeen-Roche and colleagues for HRS,(21,22) Mekli and colleagues’ for ELSA,(23) and Santos-Eggimann and colleagues’ for SHARE.(24). Please consult these for the definitive details. Briefly presented in table 1, when at least three out of five indicators are met, the person is considered frail while meeting zero to two indicators is considered together i.e. normal and pre-frail are combined as the reference; altogether making frailty a binary outcome. The indicators are exhaustion, unexpected weight loss, weakness, low energy and slowness.

**Table 1.** Variables for five indicators of frailty phenotype in previous empirical studies of Fried, Bandeen-Roche and colleagues (HRS), Mekli and colleagues (ELSA), Santos-Eggimann and colleagues (SHARE).(21–24). CESD: Center for Epidemiologic Study Depression scale.

| **Indicators** | **HRS** | **ELSA** | **SHARE** |
| --- | --- | --- | --- |
| **Exhaustion** | CESD item | CESD item | Self-report |
| **Weight loss** | Body mass index | Body mass index | Self-report of appetite |
| **Weakness** | Grip strength | Grip strength | Grip strength |
| **Low energy** | Self-report of physical activity | Self-report of physical activity | Self-report of physical activity |
| **Slowness** | Gait speed | Gait speed | Self-report of two mobility problems |

### Childhood information

In HRS, ELSA and SHARE participants were asked prospectively about their health and retrospectively about their childhood conditions up to eight decades earlier. Measures of childhood poverty are based on retrospective reports of adults aged 50–95, raising concern about recall error. For example, in 2008 some 2500 Britons aged 50 were asked to recall the numbers of rooms and people in their homes when they were eleven (to assess overcrowding). Only one in three got both numbers right (their mothers gave the correct answers when visited 39 years earlier – the families are Britain’s National Child Development Study – NCDS cohort).(1,2) Here the cross-country samples have an average age of 66. It is scarcely plausible that the NCDS peers of sixteen years older achieved perfect recall. Now recent studies have responded to the concern of recall error by devising latent construct for childhood poverty.(1–7) Following these, the latent class of childhood poverty, derived from sets of childhood condition indicators, is used as the key risk factor in old age health.

The empirical literature on cross-country comparison offers another building block to enable comparison of indicators despite less than exact sameness e.g. on depression between America and Europe as well as on allostatic load between America and Britain. Courtin and colleagues, for instance, compare two measures of depression: CESD and EuroD.(25) They found that although these measures differ in their tendency to assign caseness (CESD assigned more cases of depression), these are nevertheless comparable in risk factor associations. The focus can therefore be placed on the associations rather than abandoning comparisons altogether because of any difference in tendency. Another study similarly used different sets of indicators for allostatic load on both sides of the Atlantic, motivated by the same principle of focusing not on the indicators but on the risk factor associations.(26) In fact, as argued in presenting the method above, a latent construct to assess childhood poverty is precisely at home with different contexts or different sets of indicators.(1,2) Last, following the empirical literature studying America, Britain and Europe, all analyses control for differences between countries by including country fixed effects.(3,4)

ELSA collected life history in wave 3, while SHARE collected childhood information in its life history waves (3 and 7 for those who did not provide it the first time). The ELSA and SHARE indicators have been described elsewhere.(2,3,27,28) HRS collected childhood information in the core interview at each wave but, crucially, this is not closely comparable with indicators from the other side of the Atlantic, instead opting to elaborate on financial situation. This information is augmented with the ad hoc Life History Mail Survey 2017. Although ageing studies in America, Britain and Europe have close family resemblance, the differences are worth listing. Table 2 lists variables collected in each study, noting the emphasis in HRS on financial information, used to derive latent class of childhood poverty in a few studies.(2,3,27,28)

**Table 2.** Retrospective childhood information in HRS, ELSA and SHARE.

| Country | HRS | ELSA | SHARE |
| --- | --- | --- | --- |
| **Variable** | Number of rooms | Number of rooms | Number of rooms |
|  | Number of people | Number of people | Number of people |
|  | Number of books | Number of books | Number of books |
|  | Ever experienced:<br>being financially poor had<br>to move or be helped<br>because of financial<br>difficulty, had to live with<br>grandparents, father<br>unemployed | Physical facilities:<br>indoor toilet, hot and<br>cold running water,<br>central heating, fixed<br>bath | Physical facilities:<br>indoor toilet, hot and<br>cold running water,<br>central heating, fixed<br>bath |

### Missing data or non-response for retrospective interviews

The analytic sample comprises prospective sample members who agreed to retrospective interviews. Not all of them did and there may be significant differences between the analytic *vs* declined sample, tested using χ^2^ for binary variable and *t* test for continuous variable (table 3). In the HRS the analytic sample has 61% females and 39% males, with 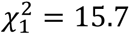 and *p* value < 0.001. In contrast, both in SHARE and ELSA analytic samples females are no more likely to agree to retrospective interviews. Meanwhile in the HRS the analytic sample has older members and this difference is significant while in SHARE and ELSA the opposite is true while also significant.

**Table 3.** Missing data or non-response patterns across analytic *vs* declined sample in HRS, SHARE and ELSA.

| Survey | Variable | Declined sample | Analytic sample |
| --- | --- | --- | --- |
| **HRS** | Female | 58% | 61% |
|  | Male | 42% | 39% |
| | | $\chi^2_1 = 15.7$ | $p < 0.001$ |
| | Mean age | 64.6 year | 68.8 year<br>$p < 0.001$ |
| **ELSA** | Female | 56% | 55% |
|  | Male | 44% | 45% |
| | | $\chi^2_1 = 2.0$ | $p = 0.160$ |
| | Mean age | 65.1 year | 64.4 year<br>$p = 0.022$ |
| **SHARE** | Female | 56% | 58% |
|  | Male | 44% | 42% |
| | | $\chi^2_1 = 1.9$ | $p = 0.165$ |
| | Mean age | 72.2 year | 67.1 year<br>$p < 0.001$ |

To test the life course shaping hypothesis, the latent class of childhood poverty was matched with frailty outcome in 2020/2021. A set of confounders or social determinants for adjustment is included as standard in the empirical literature on ageing including age, sex, youth illness, education, marital status, father/parent’s occupation, wealth, ethnicity and country (2,5,29,30) Following the literature, fixed effects model was estimated and standard error correction was applied because the key exposure of childhood poverty is an estimated latent class.(3,31) For estimation, Latent GOLD syntax version 6 was used.(32)

## Results

Table 4 collects features of the analytic sample which has 57% female, one in five was childhood poor and mean age 66.3 year. Females reported more frailty (11.7%). The bottom part shows the numbers contributed to the analytic sample by each country. The binary dependent variable frailty is split into frail and combined pre- and non-frail, with the combination as the reference level. Times ill in youth is non-negligible. Importantly, one in five experienced poverty in childhood.

**Table 4.**
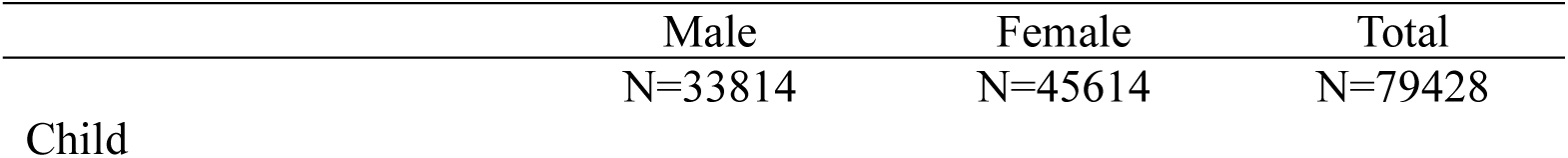
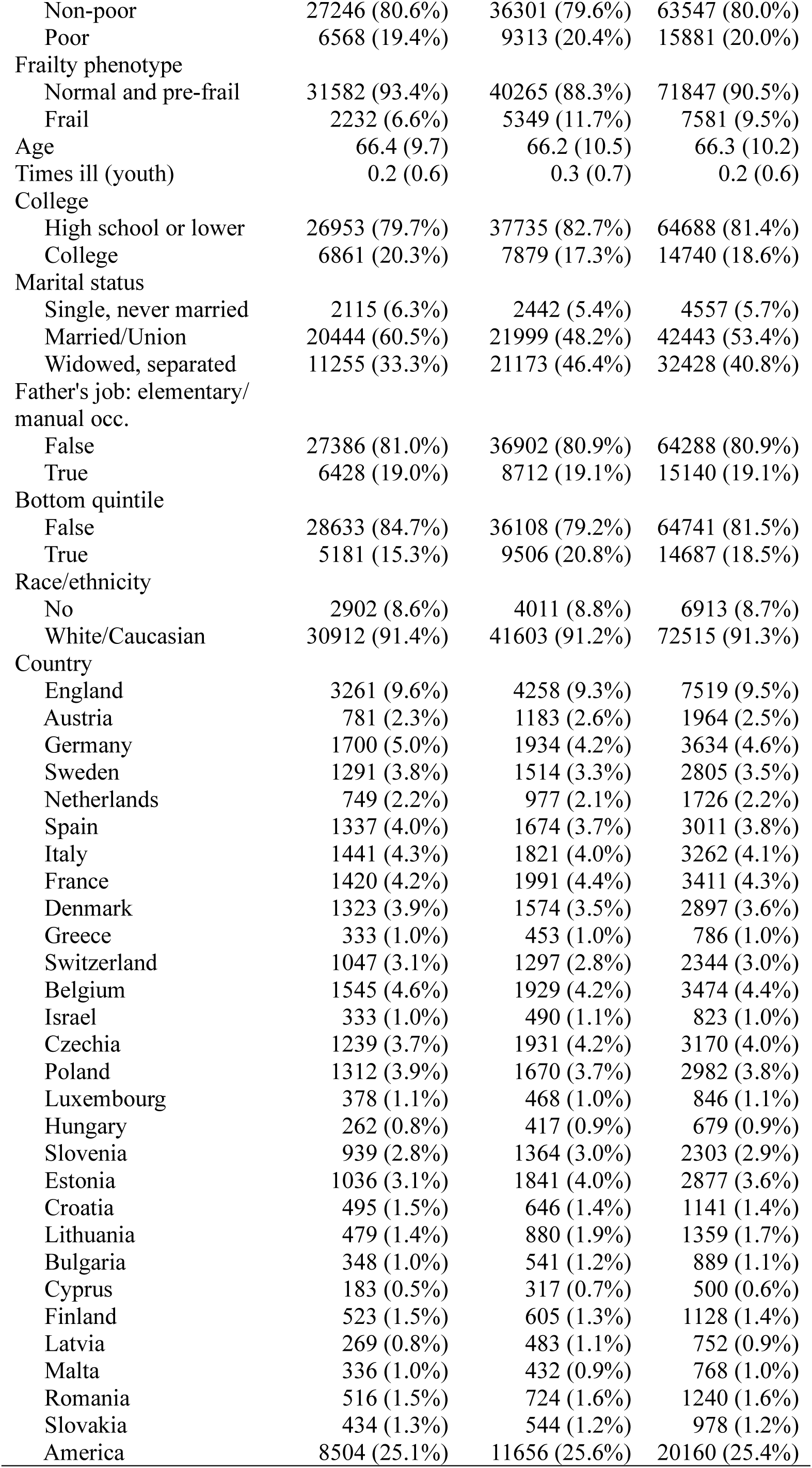
Summary features of the analytic sample with categorical variables in numbers (percentages) and continuous variables in means (standard deviations). Sources HRS, ELSA and SHARE.

The probit model coefficients explaining frailty are collected in table 5 which shows that the childhood poor report significantly higher probabilities of frailty (0.1097 ± 0.0169, *p* value ≤0.0001). In America, Britain and Europe growing up in poverty goes with growing old in frailty (coefficient 0.1097, *p* ≤0.0001), females report higher probability of being frail (0.3051, *p*≤0.0001) and the probability of frailty increases as people age (0.0398, *p*≤0.0001). Briefly, the other covariates suggest that college education (compared to high school or lower) associates with lower risks of being frail (−0.2540, *p*≤0.0001). Importantly, reflecting the social determinants framework above, youth illness is significant and in the expected direction (0.2334, *p*≤0.0001). Being in the bottom third of wealth distribution associates with higher risks of frailty, so does having a father with manual or elementary occupation. This leaves childhood poverty coefficient to be interpreted as a fully adjusted association.

**Table 5.** Probit coefficients explaining frailty in HRS, ELSA and SHARE. Constant includes the reference country Britain.

| Variable | Coefficient | Standard error | 90% conf. | interval | P value |
| --- | --- | --- | --- | --- | --- |
| Constant | -5.2006 | 0.0700 | -5.3378 | -5.0634 | $\leq 0.0001$ |
| Childhood poor | 0.1097 | 0.0169 | 0.0766 | 0.1428 | $\leq 0.0001$ |
| Sex, Female | 0.3051 | 0.0152 | 0.2753 | 0.3349 | $\leq 0.0001$ |
| Age | 0.0398 | 0.0008 | 0.0382 | 0.0414 | $\leq 0.0001$ |
| College | -0.2540 | 0.0212 | -0.2956 | -0.2124 | $\leq 0.0001$ |
| Single, never married | 0.0098 | 0.0353 | -0.0594 | 0.0790 | 0.78 |
| Married/union | -0.0914 | 0.0174 | -0.1255 | -0.0573 | $\leq 0.0001$ |
| Youth illness | 0.2334 | 0.0090 | 0.2158 | 0.2510 | $\leq 0.0001$ |
| Father had manual occ. | 0.0346 | 0.0180 | -0.0007 | 0.0699 | 0.055 |
| Bottom wealth tertile | 0.1816 | 0.0203 | 0.1418 | 0.2214 | $\leq 0.0001$ |
| Country ref: England |  |  |  |  |  |
| Austria | 1.0666 | 0.0463 | 0.9759 | 1.1573 | $\leq 0.0001$ |
| Germany | 0.9874 | 0.0425 | 0.9041 | 1.0707 | $\leq 0.0001$ |
| Sweden | 0.7525 | 0.0465 | 0.6614 | 0.8436 | $\leq 0.0001$ |
| Netherlands | 0.8745 | 0.0567 | 0.7634 | 0.9856 | $\leq 0.0001$ |
| Spain | 1.2681 | 0.0401 | 1.1895 | 1.3467 | $\leq 0.0001$ |
| Italy | 1.3377 | 0.0402 | 1.2589 | 1.4165 | $\leq 0.0001$ |
| France | 1.1892 | 0.0398 | 1.1112 | 1.2672 | $\leq 0.0001$ |
| Denmark | 0.8807 | 0.0475 | 0.7876 | 0.9738 | $\leq 0.0001$ |
| Greece | 1.3729 | 0.0614 | 1.2526 | 1.4932 | $\leq 0.0001$ |
| Switzerland | 0.6484 | 0.0530 | 0.5445 | 0.7523 | $\leq 0.0001$ |
| Belgium | 1.1513 | 0.0407 | 1.0715 | 1.2311 | $\leq 0.0001$ |
| Israel | 1.4092 | 0.0588 | 1.2940 | 1.5244 | $\leq 0.0001$ |
| Czechia | 0.9814 | 0.0423 | 0.8985 | 1.0643 | $\leq 0.0001$ |
| Poland | 1.3533 | 0.0435 | 1.2680 | 1.4386 | $\leq 0.0001$ |
| Luxembourg | 0.8424 | 0.0753 | 0.6948 | 0.9900 | $\leq 0.0001$ |
| Hungary | 1.3522 | 0.0653 | 1.2242 | 1.4802 | $\leq 0.0001$ |
| Slovenia | 0.9299 | 0.0460 | 0.8397 | 1.0201 | $\leq 0.0001$ |
| Estonia | 1.1239 | 0.0410 | 1.0435 | 1.2043 | $\leq 0.0001$ |
| Croatia | 1.1975 | 0.0565 | 1.0868 | 1.3082 | $\leq 0.0001$ |
| Lithuania | 1.1440 | 0.0547 | 1.0368 | 1.2512 | $\leq 0.0001$ |
| Bulgaria | 1.3211 | 0.0594 | 1.2047 | 1.4375 | $\leq 0.0001$ |
| Cyprus | 1.2528 | 0.0720 | 1.1117 | 1.3939 | $\leq 0.0001$ |
| Finland | 0.7931 | 0.0687 | 0.6584 | 0.9278 | $\leq 0.0001$ |
| Latvia | 0.9777 | 0.0699 | 0.8407 | 1.1147 | $\leq 0.0001$ |
| Malta | 1.1812 | 0.0668 | 1.0503 | 1.3121 | $\leq 0.0001$ |
| Romania | 1.3181 | 0.0554 | 1.2095 | 1.4267 | $\leq 0.0001$ |
| Slovakia | 1.5224 | 0.0591 | 1.4066 | 1.6382 | $\leq 0.0001$ |
| America | 0.3587 | 0.0404 | 0.2795 | 0.4379 | $\leq 0.0001$ |

To bring this forward, for each of the 29 countries the predicted probabilities of frailty are plotted as distinguished by childhood poverty over age 70 to 90. This shows at a glance how childhood poverty shapes frailty in old age across a wide range of history and health systems of rich countries.

**Figure 1.**
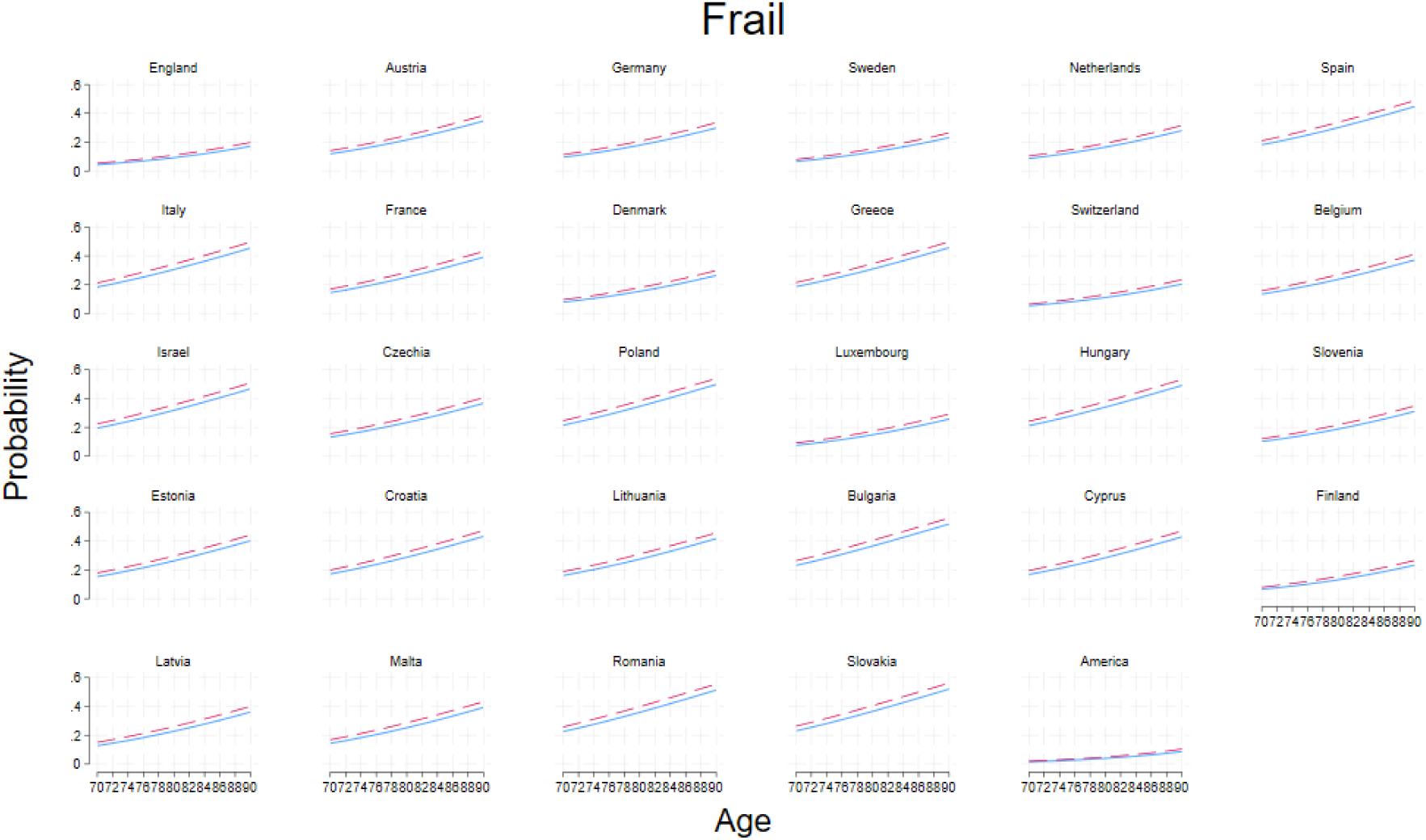
Probabilities of Frail among the childhood poor (dash) and non-poor (solid) in older people aged 70 to 90 years in Britain, Europe and America based on models in table 3 where all covariates are set at the sample averages. Analysis of HRS, ELSA and SHARE.

The trellis plot reveals several key patterns. First, age is an important risk factor though it works differently in different countries, validating the fixed effect design. For example, compare the levels and slopes of Slovakia and America (the last two panes). Second, childhood poverty puts people at a disadvantage in old age, putting the dashed lines always above the solid lines. This is statistically significant as shown in Table 5. Third, the regional patterns of Europe are pronounced (Northern, Southern and Eastern Europe). This can be seen for Sweden, Finland and Denmark as a group, Spain, Italy and Greece as another, and Croatia, Romanie and Slovakia as yet another. Together, the variation is marked, preventing one single summary pattern from representing all high income countries.

To check the robustness of these results, two sensitivity analyses were conducted replacing the fixed effect design with a random effect one, effectively taking these countries as randomly drawn from a population of rich countries, then stratifying by sex. The results above stand (table 6 and 7).

**Table 6.** Sensitivity analysis: Probit coefficients with random effects explaining frailty in America (HRS), Britain (ELSA) and Europe (SHARE)

| Variable | Coefficient | Standard error | P value |
| --- | --- | --- | --- |
| Const | -4.1054 | 0.2494 | $\leq 0.0001$ |
| Childhood poor | 0.1509 | 0.0244 | $\leq 0.0001$ |
| Sex, Female | 0.3068 | 0.0620 | $\leq 0.0001$ |
| Age | 0.0406 | 0.0032 | $\leq 0.0001$ |
| College | -0.2609 | 0.0242 | $\leq 0.0001$ |
| Single, never married | 0.0139 | 0.0290 | 0.63 |
| Married/union | -0.0848 | 0.0223 | $\leq 0.0001$ |
| Youth illness | 0.2386 | 0.0114 | $\leq 0.0001$ |
| Father had manual occ. | 0.0483 | 0.0298 | 0.10 |
| Bottom wealth tertile | 0.1886 | 0.0559 | 0.0007 |
| Random effect variance | 0.0989 | 0.0113 | $\leq 0.0001$ |

**Table 7.**
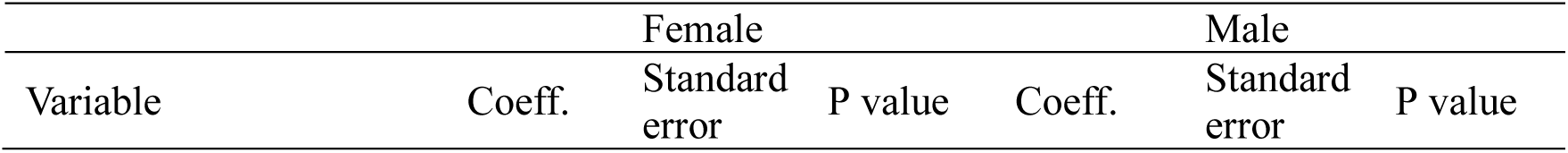
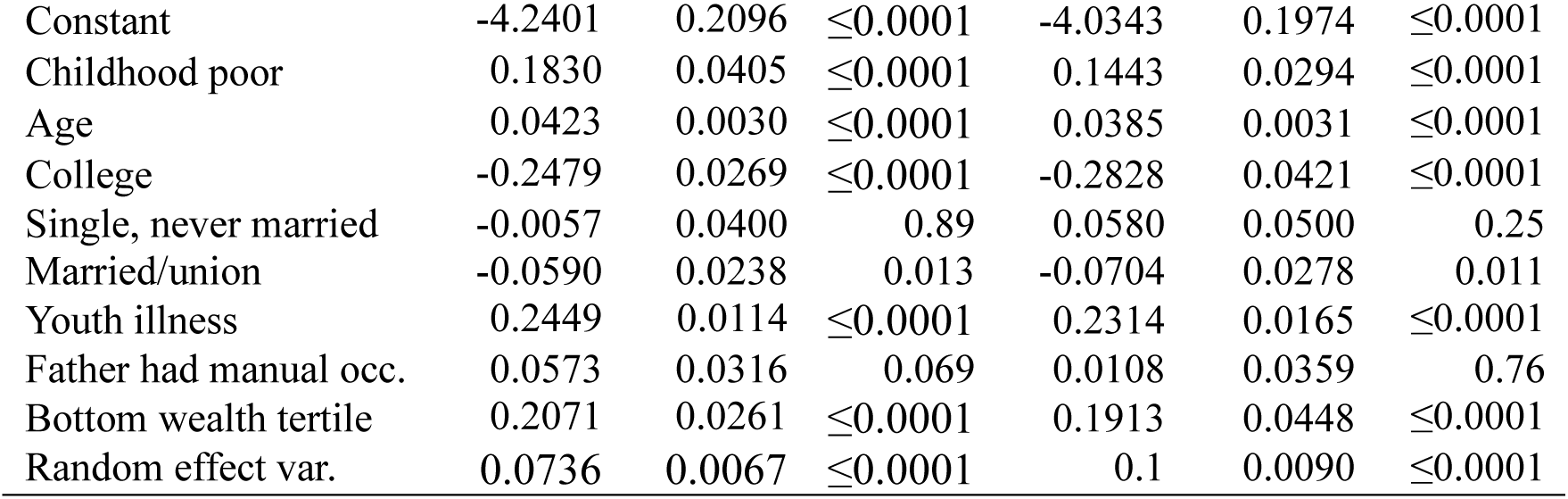
Sensitivity analysis: Probit coefficients with random effects and stratified by sex, explaining frailty in America (HRS), Britain (ELSA) and Europe (SHARE)

## Discussion and conclusion

Evidence is accumulating and compelling that the life course shaping of health reaches into old age. Childhood poverty continues to shape the health of older people around the world even in rich nations across wide outcomes of ageing including frailty. This is the first evidence in 29 nationwide representative populations of how childhood poverty plays a role in frailty variations in older adults. This operates in diverse health systems: in a national health system (UK), a largely private health system (US) and everything in between (27 European countries). The UK NHS is more than 70 years old and has accompanied Britons in the study cohorts throughout their life courses. Using latent construct to recover child poverty status from sets of error-laced indicators, the analysis shows important associations between childhood poverty and frailty that are consistently observed across 29 countries enriching our appreciation of the life course shaping of health in old age.

What mechanism lies behind this shaping is an important question. I have tried to offer an epigenetic explanation and evidence for the role of childhood poverty as a risk factor.(3) Poverty in childhood is posited to induce epigenetic change through increasing methylation rates which leaves a mark that lasts until old age. Children with similar genetic make-up may nevertheless differ in their phenotypic expression, hence organ function, because their childhood conditions differ in ways that change adverse methylation rate in one child more than in another. That childhood lasts a lifetime, we knew. Now we know it also shapes the chance of frailty syndrome.

### Limitations and strengths

There are limits to what can be learned from this work. First, its framework is not a full structural framework,(4) although this has precedents.(2,5,33) That alternative has encouraged the authors to suggest that were more covariates considered, the association with childhood poverty would be entirely weakened. This suggestion is of course an eminently empirical question. The imperative remains that a fuller framework should be considered when comparable information on youth and adult conditions in all these countries are available. Second, this work is also limited in its focus on material lack in childhood, lacking psychological variables such as parent-child bonding or nurturing relationships. Cultivating psychological development is of course core to the life course stage of raising children. Third, by including fixed effects for countries this work controls for unobserved differences between countries such as their histories and health systems. Despite precedent in using country fixed effects, this may be considered insufficient and thus constitutes another limitation. Country by country analyses for these 29 countries are deferred to future monographs. Last, a key limitation arises from the design being observational rather than a randomised study (to childhood material intervention and to control), preventing causal interpretation of these results. The childhood poor may differ from the non-poor in ways not observed. Moreover, some of the childhood poor may not have survived to take part in these longitudinal ageing studies. Although this survivor effect may suggest a direction of bias, its magnitude is unknown. Last, as noted above the analytic sample here differs in many ways from those who declined to give childhood information. But across 29 countries these are not entirely systematic.

The strengths were mentioned throughout but three need emphasis. First, this is the first analysis to study 29 countries in the family of Health and Retirement Study, with closely comparable key exposure and health outcomes that speak directly to the UN Decade of Healthy Ageing. The geographic reach of the sample as well as the variety of health systems it encompasses, together strengthen the generalisation of the results to other rich nations.

Second, all the parallel surveys are ongoing, making this work a unique pad to launch future works on the consequences of child poverty in old age, on age trajectories of myriad health outcomes in ageing populations in rich nations. Lastly, it shows what can be achieved with a clear framework and a robust method applied to a family of ageing studies.

### Reflection on recent literature

These findings echo recent studies in different countries though not all.(1–3,5–7,27,28) While consensus has not yet arrived, the evidence is largely supportive of a life course shaping of frailty. The contrast with the new findings here can arise from the outcome studied or the special nature of the sample. Here nationally representative samples are used, comparable across 29 countries. Further, fixed effects capturing idiosyncratic country effects such as unique health system and history, are included. All agree in showing that the childhood poor have more frailty in old age.

But a few studies on the life course shaping of health in older ages have reported different results, which may be due to variations in variable construction, outcomes, methods, health systems and broader structures of societies. Vable and coauthors in America suggest that once socioeconomic mobility in adulthood is considered, no direct association remains between childhood economic status and old age health.(7) On this side of the Atlantic, studies in Sweden and Europe have also shown varying associations between poverty or economic status in childhood with old age health.(4–6) But the construct of childhood conditions vary. Lennartsson and colleagues depart from the usual practice of ignoring recall bias by using latent construct of childhood poverty, in fact a full structural model to explain old age health indicated by pain, fatigue and breathing difficulty in Sweden; meanwhile Pakpahan and colleagues also used a structural model to explain self-rated health in old age in 13 European countries.(4,5)

In Sweden the authors suggest that childhood poverty is no longer binding on old age health once adult conditions are considered. This may be explained in two ways. The extensive welfare state of Sweden has been known to deliver exceptional health service to its older population. Our own work comparing 17 countries in Europe shows that Sweden has managed to break the link between economic position and sensory impairment.(34) Lennartsson and colleagues may have found a manifestation of Sweden’s exceptionalism in ameliorating pain, fatigue and breathing difficulty. But this can sit with the results above: on frailty childhood poverty continues to matter.(5) Sweden’s exceptionalism may not have succeeded in breaking all the links between childhood poverty and the spectrum of health in old age. If indeed the mechanism involves epigenetic changes, amelioration is possible but perhaps the chance of complete elimination is slim.

### Future research and policy including the UN Decade of Healthy Ageing

In England the Chief Medical Officer recently reported on critical plans to ensure healthy ageing in the country.(10) The report entitled “Health in an Ageing Society” emphasises within-country spatial disparity in frailty, in particular showing older adult with greater need of frailty treatment reside in coastal and rural areas which in turn presents specific extra challenges. The report reproduced a map of the extent of spatial disparity in frailty which we produce to highlight this aspect of frailty challenge that needs more investigation in any country.(9)

More broadly, the multi-country evidence laid here raises several implications for research and global health policy. First, retrospective childhood information can be more fruitfully examined by recognising that this information is laced with error. Childhood poverty, a latent construct indicated by several items of recalled information, can be obtained as a latent class.(1–3) Second, this strong result from many countries speaks directly to global health policy such as the [UN Decade of Healthy Ageing](https://www.who.int/initiatives/decade-of-healthy-ageing). The initiative has missed an opportunity to grab the life course shaping of health in older ages around the world. If rich countries with their advanced health systems have experienced this life course shaping of health, the low and middle income countries are also likely to carry the long reach of childhood poverty. The 29 countries in this study are high income countries; those who grew rich before growing older. Other countries are in a decidedly more difficult predicament of growing old before growing rich e.g. India and China. The WHO observed that the countries with the difficult predicament are also growing old faster: what took France more than a century will take China only a few decades.(35) These countries should be the focus of future research on the life course shaping of health in older ages. With the higher prevalence of childhood poverty in low and middle income countries and its long arm reaching into old age as evinced here, the need for research in low and middle countries is urgent.

In conclusion, childhood poverty reaches long into old age, risking frailty in older ages. While more knowledge is needed, especially from low and middle income countries on how epigenetic changes operate, strong evidence is at hand to help secure a decade of healthy ageing by proving the necessity of eliminating child poverty.

## Data Availability

All data used are available online at https://www.elsa-project.ac.uk, https://hrs.isr.umich.edu, DOI: 10.6103/SHARE.w3.800, 10.6103/SHARE.w7.800, 10.6103/SHARE.w9ca800

https://hrs.isr.umich.edu

https://doi.org/10.6103/SHARE.w3.800

https://doi.org/10.6103/SHARE.w7.800

https://doi.org/10.6103/SHARE.w9ca800

## Ethical review

The University of Manchester exempted the investigation from full ethical review as it uses publicly available deidentified secondary datasets.

## STROBE list

STROBE list is attached.

## Acknowledgement

I thank the participants in 29 countries for providing information, time and in many visits blood biomarkers too, as well as the generous funding bodies over many decades. **HRS**: The HRS (Health and Retirement Study) is sponsored by the National Institute on Aging (grant number NIA U01AG009740) and is conducted by the University of Michigan. **ELSA**: The English Longitudinal Study of Ageing was developed by a team of researchers based at University College London, NatCen Social Research, the Institute for Fiscal Studies, the University of Manchester and the University of East Anglia. The data were collected by NatCen Social Research. The funding is currently provided by the National Institute on Aging in the US (grant numbers: 2RO1AG7644 and 2RO1AG017644-01A1), and a consortium of UK government departments coordinated by the National Institute for Health Research. **SHARE**: This paper uses data from SHARE Waves 3, 7 and 9 (DOIs: 10.6103/SHARE.w3.800, 10.6103/SHARE.w7.800, 10.6103/SHARE.w9ca800) see Börsch-Supan et al. (2013) for methodological details.(1) The SHARE data collection has been funded by the European Commission, DG RTD through FP5 (QLK6-CT-2001-00360), FP6 (SHARE-I3: RII-CT-2006-062193, COMPARE: CIT5-CT-2005-028857, SHARELIFE: CIT4-CT-2006-028812), FP7 (SHARE-PREP: GA N°211909, SHARE-LEAP: GA N°227822, SHARE M4: GA N°261982, DASISH: GA N°283646) and Horizon 2020 (SHARE-DEV3: GA N°676536, SHARE-COHESION: GA N°870628, SERISS: GA N°654221, SSHOC: GA N°823782, SHARE-COVID19: GA N°101015924) and by DG Employment, Social Affairs & Inclusion through VS 2015/0195, VS 2016/0135, VS 2018/0285, VS 2019/0332, and VS 2020/0313. Additional funding from the German Ministry of Education and Research, the Max Planck Society for the Advancement of Science, the U.S. National Institute on Aging (U01_AG09740-13S2, P01_AG005842, P01_AG08291, P30_AG12815, R21_AG025169, Y1-AG-4553-01, IAG_BSR06-11, OGHA_04-064, HHSN271201300071C, RAG052527A) and from various national funding sources is gratefully acknowledged (see www.share-project.org).

